# The Hemodynamic Burden of Coronary Artery Tortuosity in Takotsubo Syndrome: A Closer Look at Coronary Flow and Myocardial Stunning

**DOI:** 10.64898/2026.01.26.26344895

**Authors:** Arthur Alencar, Jash Sash, Sumair Ozair, Christopher Railwah, Barry Bertolet

## Abstract

**Background:** Coronary artery tortuosity (CAT) is often viewed as a benign angiographic finding; however, emerging evidence suggests its potential hemodynamic significance, particularly in non-atherosclerotic cardiomyopathies such as Takotsubo syndrome (TS).

**Objectives:** This study aimed to investigate the prevalence and hemodynamic implications of CAT in patients diagnosed with Takotsubo cardiomyopathy (TCM) and to evaluate the association between the severity of tortuosity and myocardial injury markers, recovery of ventricular function, and other clinical variables.

**Methods:** A retrospective review of 100 patients with TCM from the Baptist Memorial Hospital network (2015–2025) was conducted. Tortuosity severity was quantified using angiographic criteria per Eleid et al. (2014). Associations between CAT and biochemical or echocardiographic parameters were evaluated using multiple linear regression and non-parametric tests.

**Results:** CAT was highly prevalent (85.1%) in this TCM cohort, with a mean tortuosity index of 3.26—significantly higher than in general angiography populations. No significant correlations were found between tortuosity severity and peak troponin levels (p = .588) or ejection fraction (EF) at presentation (p = .820). Full EF recovery (55–65%) at ≥3 months occurred in 70.7% of patients and was not significantly associated with prior cardiomyopathy, coronary artery tortuosity index or baseline troponin levels.

**Conclusions:** CAT appears markedly more prevalent among patients with TCM, although its severity does not correlate with biomarker elevation or EF recovery. These findings suggest that coronary tortuosity may contribute to the hemodynamic environment predisposing to TS, without directly determining the extent of myocardial dysfunction or recovery.

## Introduction

Coronary artery tortuosity (CAT) is a relatively common finding on angiography and was traditionally considered a benign incidental feature, primarily noted when proximal to a lesion, where it could complicate percutaneous coronary intervention. CAT is typically defined as three or more consecutive 45°–90° curves in a major epicardial artery, and it is considered severe when two or more 180° curves are present.

Recent studies have associated CAT with non-atherosclerotic cardiomyopathy. Estrada et al. [1] reported a statistically significant association between CAT and myocardial ischemia in a retrospective cohort. Similarly, studies have identified myocardial ischemia in patients without obstructive coronary artery disease [2], suggesting that unrecognized structural or hemodynamic factors, such as tortuosity, may contribute to myocardial dysfunction.

Computer simulations have demonstrated that increased tortuosity correlates linearly with decreased perfusion pressure [4,5]. Takotsubo cardiomyopathy (TCM), which presents as acute coronary syndrome (ACS) with transient LV dysfunction and elevated cardiac biomarkers in the absence of obstructive lesions [6], may share underlying hemodynamic mechanisms with CAT.

This study aims to determine the association between CAT and Takotsubo cardiomyopathy, examining the relationship between tortuosity severity and myocardial injury, functional recovery, and other clinical variables.

## Methods

### Study Population

This retrospective study included 100 patients from the Baptist Memorial Hospital network (East Arkansas, North Mississippi, and West Tennessee) diagnosed with TCM between 2015 and 2025.

Inclusion criteria required a cardiologist-confirmed diagnosis of TCM with echocardiographic findings within 24 hours of coronary angiography showing no significant atherosclerotic disease. Patients with incomplete echocardiographic or angiographic data were excluded.

Collected variables included initial and peak troponin levels, ejection fraction (EF) at diagnosis, Pro-BNP, and follow-up EF (≥3 months). Additional data encompassed history of fibromuscular dysplasia, hypertension (with stage), smoking status, and use of guideline-directed medical therapy (GDMT) at discharge (SGLT2 inhibitors, ACE/ARB/ARNI, β-blockers, and aldosterone antagonists). Angiograms were reviewed by investigators and cardiologists, who were not blinded to patient data.

### Tortuosity Definitions

Following Eleid et al. [7], CAT was defined as ≥3 consecutive curvatures ranging 90°– 180° in a major epicardial coronary vessel (≥2 mm diameter).

Severe: ≥2 consecutive curvatures ≥180° in a ≥2 mm vessel

Moderate: ≥3 consecutive 90°–180° curvatures in ≥2 mm vessel (not meeting severe criteria)

Mild: ≥3 consecutive 45°–90° curvatures in ≥2 mm vessel, or ≥3 90°–180° curvatures in an artery <2 mm

Each major epicardial artery (LAD, LCx, RCA) was assigned a score (0–3), producing a composite Tortuosity Severity Index.

### Statistical Methods

Continuous variables were reported as means ± SD and categorical variables as frequencies or percentages. The prevalence of CAT was defined as the proportion with a tortuosity index >0. Associations between tortuosity and clinical or biochemical parameters were analyzed using multiple linear regression, with independent variables including age, troponin (initial and peak), Pro-BNP, EF, smoking status, hypertension stage, and GDMT.

Statistical significance was set at p < .05, while p values between .05 and .10 were interpreted as trends toward significance. Analyses were conducted in Python using pandas and statsmodels.

### Limitations

This study has several important limitations. First, its retrospective design inherently introduces selection and information bias, as data availability and documentation quality may vary across patients and over the study period. Second, coronary tortuosity assessment, while based on standardized angiographic criteria, retains an element of subjective interpretation, which may lead to interobserver variability. This limitation is amplified by the fact that angiographic reviewers were **not blinded** to clinical data, potentially introducing observer bias in tortuosity grading.

## Results

### Prevalence of Coronary Artery Tortuosity

CAT was present in 85.1% of the cohort, with a mean tortuosity index of 3.26— substantially higher than the prevalence observed in general angiographic populations, where rates average 45.2% in women and 19.7% in men (Chiha et al., 2016). Among female patients in this study, CAT prevalence reached 85.7%, reinforcing its potential role as a contributing risk factor for TCM (OR = 7.27; 95% CI, 3.0–17.6; p < .0001). Notably, patients with SCAD demonstrated a higher mean tortuosity score (4.41 ± 1.73) compared with those without SCAD (2.33 ± 1.49) [4], suggesting a shared vascular vulnerability across these two conditions.

### Baseline Characteristics and Association Between CAT and Clinical Variables

Baseline continuous clinical variables stratified by the presence of any coronary tortuosity (index > 0) are presented in Table 1. No statistically significant differences were observed for age, troponin values, pro-BNP, or hypertension stage between patients with and without tortuosity.

### Association Between CAT Index and Troponin Levels

No significant association was found between tortuosity index and troponin levels (initial: p = .616; peak: p = .588). Given that TCM primarily involves myocardial stunning rather than necrosis, biomarker elevation did not correlate with tortuosity severity.

**Table 1.**
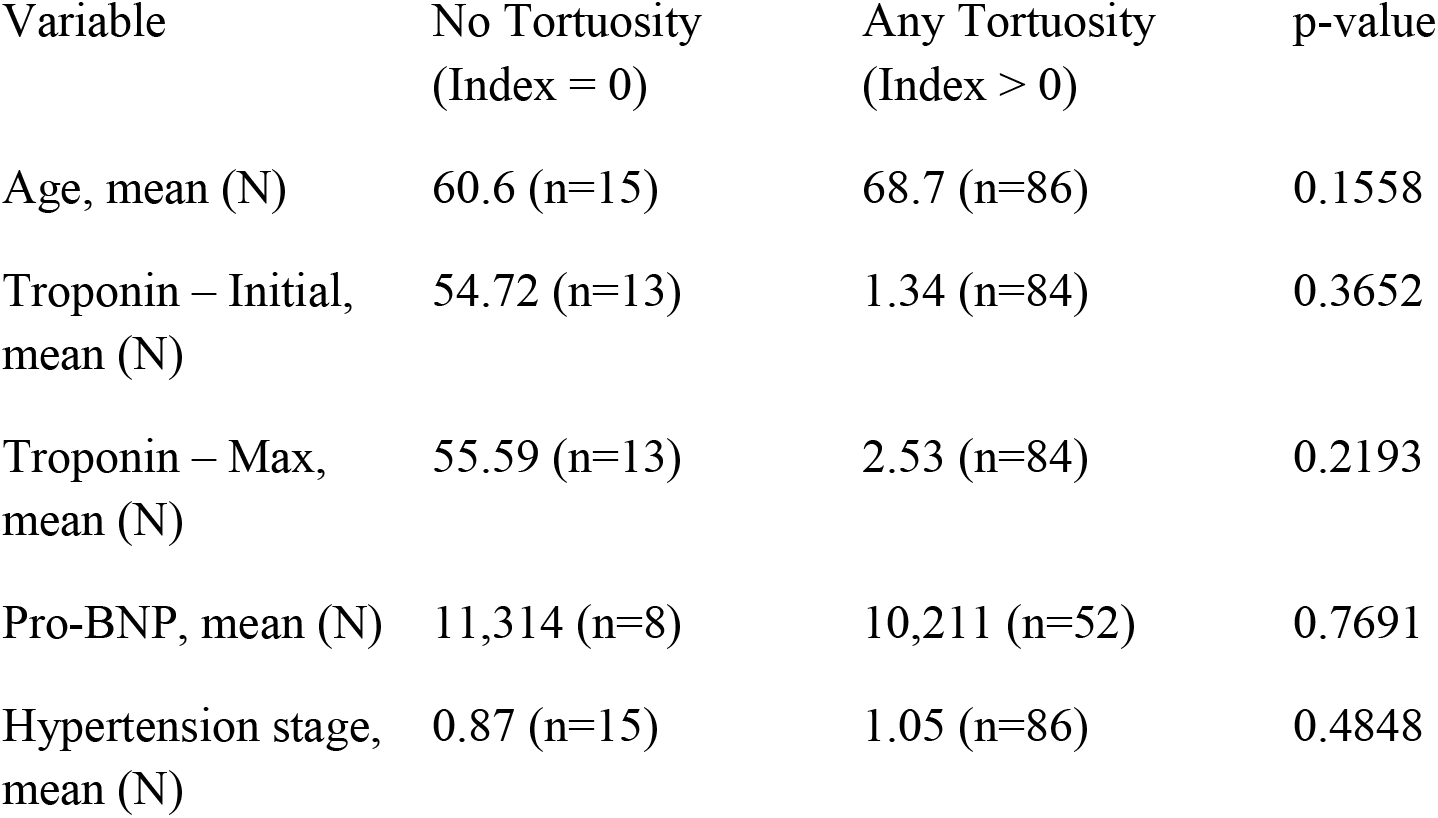
Continuous variables by presence of any coronary artery tortuosity.

### Association Between Tortuosity Index and Ejection Fraction

Baseline EF at time of diagnosis averaged 30.0%, with no significant relationship between EF and tortuosity (β = –0.003, p = .820). Even though TCM is primarily thought of as a condition related to LV dysfunction, prior studies indicate that right ventricular involvement may be more common than expected, which would confound EF measurement in TCM [19].

### Association Between Tortuosity and Hypertension Stage

Although CAT has been linked to hypertension, this study found no significant relationship between hypertension stage and tortuosity (β = +0.481, p = .063). The near-significant value suggests the need for larger, more comprehensive studies. The definition of hypertension stage used was ACC/AHA 2017 independent of current hypertensive use.

### Observed Recovery of Left Ventricular Function

Full LV recovery—defined as EF between 55–65% on follow-up echocardiogram ≥3 months—was observed in 70.7% of patients. Recovery according to presence or absence of tortuosity is summarized in Table 2.

**Table 2.** EF recovery vs any tortuosity.

| EF Recovery Status | No Tortuosity | Any Tortuosity | p-value |
| --- | --- | --- | --- |
| Recovered | 5 | 48 |  |
| Not recovered | 5 | 17 |  |
| Total | 10 | 65 | 0.2425 |

**Table 3.** EF recovery vs prior cardiomyopathy.

| EF Recovery Status | No Prior CM | Prior CM | p-value |
| --- | --- | --- | --- |
| Recovered | 48 | 5 |  |
| Not recovered | 18 | 4 |  |
| Total | 66 | 9 | 0.5021 |

### Impact of Prior Cardiomyopathy on Recovery

The impact of prior cardiomyopathy on EF recovery is summarized in Table 4. Prior cardiomyopathy was not significantly associated with reduced likelihood of EF normalization.

**Table 4.** Logistic regression for EF recovery.

| Predictor | Coefficient ($\beta$) | Odds Ratio (OR) | p-value |
| --- | --- | --- | --- |
| Intercept | 0.0781 | 1.08 | 0.903 |
| Any tortuosity | 1.0750 | 2.93 | 0.124 |
| Prior cardiomyopathy | -0.8120 | 0.44 | 0.271 |

### Multivariable Logistic Regression for EF Recovery

A multivariable logistic regression model including any tortuosity and prior cardiomyopathy as predictors of EF recovery did not identify statistically significant associations. Model coefficients and odds ratios are presented in Table 4.

## Discussion

Multiple mechanisms have been proposed for LV dysfunction in Takotsubo syndrome. We aim to discuss the main mechanisms and shed new light on the most promising explanations.

### Catecholamine-Induced Cardiomyopathy

The association between elevated norepinephrine and epinephrine levels and the development of myocardial ischemia is well established in the literature [6]. These observations suggest that catecholamine excess may contribute, at least in part, to myocardial dysfunction through direct myocyte injury. However, this mechanism alone does not fully account for the clinical features observed in known catecholamine-mediated cardiomyopathies such as phaeochromocytoma-induced cardiomyopathy [8]. In Takotsubo syndrome specifically, the absence of late gadolinium enhancement on cardiac magnetic resonance imaging indicates a lack of significant myocardial necrosis, distinguishing TS from classical forms of catecholamine-induced myocardial damage.

### Vasospasm

Another proposed mechanism for Takotsubo syndrome involves coronary vasospasm leading to transient myocardial ischemia. This hypothesis was first introduced by Dote et al. [9], who reported that 4 of the initial 5 patients exhibited vasospasm during angiography. Subsequent small studies similarly reported associations between vasospasm and TS [10,11]. Moreover, transient apical ballooning following intracoronary acetylcholine administration has been documented in patients undergoing evaluation for Takotsubo cardiomyopathy [12], further supporting a vasospastic component.

Given the high prevalence of CAT in the Takotsubo population, its potential relationship with vasospasm has also been explored. Jansen et al. [13] evaluated 228 patients—73 with no tortuosity, 114 with mild tortuosity, and 41 with moderate tortuosity— undergoing acetylcholine-provoked angiography for chest pain, and found no significant differences between groups in the likelihood of major epicardial spasm, defined as >90% reduction in vessel diameter. In contrast, McChord et al. [14] identified a higher prevalence of coronary tortuosity among patients with microvascular spasm (58%) compared with those with epicardial spasm (43%) or with negative/inconclusive acetylcholine testing (49%) (P = 0.017), suggesting that CAT may predispose specifically to microvascular spasm rather than large-vessel spasm. However, additional studies, particularly those including patients with severe tortuosity, are needed to better define this relationship.

The association between Takotsubo syndrome and microvascular spasm was further supported by Vitale et al. [15], reinforcing the concept that microvascular dysfunction— potentially exacerbated by coronary tortuosity—may play a central role in the pathogenesis of TS.

### Spontaneous Coronary Artery Dissection

The association between spontaneous coronary artery dissection (SCAD) and coronary artery tortuosity has been previously established [7], raising the question of whether SCAD and Takotsubo syndrome (TS) may share overlapping pathophysiologic mechanisms. Early reports noted that several cases initially diagnosed as Takotsubo were later reclassified as SCAD [16], as SCAD can produce a clinical and imaging presentation—including apical ballooning—that closely mimics TS. While this overlap suggests a potential unifying mechanism for a subset of cases, the relationship between SCAD and other characteristic features of TS, such as catecholamine excess, remains incompletely understood.

Gurgoglione et al. [17] observed a strong association between SCAD and emotional stressors in women, implying a possible link to catecholamine surges similar to those implicated in TS. Notably, patients with pure SCAD tend to be younger and have fewer traditional cardiovascular risk factors compared with typical TS cohorts [7,16], including the population in our study (mean age 45.3 ± 8.9 vs 65.7; Table 2). It is also relevant to note that SCAD was not documented in our patient population. In our view, these findings support the presence of a significant clinical and mechanistic overlap between SCAD and Takotsubo syndrome, suggesting that these conditions may represent different manifestations along a shared spectrum of stress-related coronary vulnerability.

## Conclusion

The characteristic findings of Takotsubo syndrome are likely driven by a combination of hemodynamic alterations affecting the coronary circulation during episodes of significant catecholamine release. Although direct catecholamine-induced myocardial injury has not been consistently demonstrated, the surge in circulating catecholamines appears to create a vulnerable microcirculatory environment that predisposes susceptible individuals to myocardial stunning. In our study, biomarker levels and presenting ejection fraction were evenly distributed across tortuosity categories, suggesting that tortuosity alone by this definition does not determine the initial severity of myocardial dysfunction. Nevertheless, a more comprehensive assessment of coronary tortuosity—paired with detailed evaluation of both left and right ventricular function in TS—may yield important mechanistic insights.

Several factors that predispose individuals to SCAD may also increase susceptibility to TS. As demonstrated by Mohsin et al. [18], patients diagnosed with SCAD were more than seven times more likely to also be diagnosed with TS. However, not all individuals with SCAD develop the classic apical ballooning pattern, highlighting the heterogeneity of this syndrome. Notably, We propose that the microvascular abnormalities associated with coronary artery tortuosity may further amplify the risk of TS, particularly in older patients and in those with traditional cardiovascular risk factors—most notably hypertension.

Future studies are needed to evaluate therapeutic strategies that target the hemodynamic and microvascular disturbances observed in this population. Such approaches have the potential not only to improve outcomes in Takotsubo syndrome but also to benefit patients with non-obstructive coronary artery disease more broadly

## Data Availability

Data can be shared upon request and internal IRB approval

